# Alternative Defibrillation Strategies for Refractory Ventricular Fibrillation: A Systematic Review and Meta-Analysis

**DOI:** 10.1101/2023.09.26.23296188

**Authors:** Benedetta Perna, Matteo Guarino, Roberto De Fazio, Ludovica Esposito, Andrea Portoraro, Federica Rossin, Francesca Remelli, Caterina Trevisan, Valeria Raparelli, Giovanni Marasco, Giovanni Barbara, Stefano Petrini, Milo Vason, Michele Domenico Spampinato, Roberto De Giorgio

**Affiliations:** Department of Translational Medicine, St. Anna University Hospital of Ferrara, University of Ferrara, Italy; Emergency Department, S. Anna University Hospital of Ferrara, Italy; Emergency Medicine, Azienda Unità Sanitaria Locale, Ferrara, Italy; Department of Medical Sciences, St. Anna University Hospital of Ferrara, University of Ferrara, Italy; IRCCS Azienda Ospedaliero Universitaria di Bologna, Bologna, Italy; Department of Medical and Surgical Sciences, University of Bologna, Bologna, Italy

**Keywords:** Alternative defibrillation strategies, cardiac arrest, double sequential external defibrillation, refractory ventricular fibrillation, resuscitation, vector-change defibrillation

## Abstract

**Background:** Cardiac arrest with refractory ventricular fibrillation (rVF) represents a dramatic medical emergency. Despite recent advances, its treatment is challenging and burdened by limited evidence. This systematic review and meta-analysis aims at establishing whether alternative defibrillation strategies (ADS), i.e. double sequential external defibrillation (DSED) or vector-change defibrillation (VCD), improve survival among patients with rVF compared to standard defibrillation (SD).

**Methods:** Randomized clinical trials (RCTs), prospective and retrospective studies were included if: (1) compared ADS with SD in rVF; (2) conducted on patients ≥ 18 years old; (3) reported survival to hospital admission. English-language papers from MEDLINE, Google Scholar, Cochrane Library, World Health Organization, EMBASE and CINAHL, published from inception to December 2022, were retrieved. The risk of bias was assessed following the National Institutes of Health Quality Assessment Tool for Observational Cohort and Cross-Sectional Studies and the revised Cochrane risk of bias tool for randomized trials, as appropriate. A random-effects meta-analysis was performed to estimate the pooled Odds Ratio (pOR) with 95% Confidence Interval (95%CI) of ADS and survival to hospital admission. Furthermore, a subgroup analysis was performed to compare SD with each type of ADS. The protocol was registered on PROSPERO (CRD42022379049).

**Results:** Eight studies (2 RCTs, 5 retrospective and 1 case-control study) were retrieved for qualitative and quantitative analyses. The study population included 1405 patients (ADS = 493 vs. SD = 912) with a pooled mean age of 61.9 ± 1.1 years; among them, 277 (19.7%) were female. The random-effect meta-analysis did not show differences in survival to hospital admission among ADS vs. SD (pOR = 1.12, 95%CI: 0.62-2.01). The subgroup analysis confirmed that neither DSED (pOR = 1.20, 95%CI: 0.56-2.58) nor VCD (pOR = 1.66, 95%CI: 0.10-27.02) were associated with improved survival to hospital admission. Main limitations were: i) few numbers of studies included with small sample size; and ii) female under-representation.

**Conclusion:** The present manuscript did not show any difference on survival to hospital admission between the considered defibrillation strategies in rVF. This result highlights the need for further *ad hoc* clinical trials assessing the actual role of ADS.

## Background

Cardiac arrest (CA) is a condition characterized by the interruption of cardiac mechanical activity in the absence of any circulation sign [1]. In 2021, out-of-hospital cardiac arrest (OHCA) has been identified by United States Emergency Medical Service (EMS) in around 150000 individuals [2], but the real incidence is unknown because most cases are not managed by EMS [3]. In-hospital cardiac arrest (IHCA) is also common, occurring almost 290000 patients in the United States every year [4]. Both conditions are burdened by poor survival rates (9.1% and 18.8% for OHCA and IHCA, respectively) and among survivors, only 7.1% and 12.9%, respectively, had good functional status [2]. Sex disparities in the epidemiology of CA have been reported. Specifically, OHCA is more common in males than females (69% *vs*. 31%). Concordantly, IHCA occurs more frequently in males (58%) with a mean age of 66 years [4,5]. CA may have two different clinical presentations: one with shockable rhythm, i.e. ventricular fibrillation (VF) or pulseless ventricular tachycardia (pVT); the other with non-shockable rhythm, i.e. asystolia or pulseless electric activity (PEA) [6]. Patients presenting with shockable rhythms have a higher survival rate than those with non-shockable ones [7]. Nevertheless, almost half of subjects with shockable rhythms may remain in refractory ventricular fibrillation (rVF), defined as VF or pVT still detectable after 3 consecutive rhythm analyses and standard defibrillations separated by 2-minute intervals [8]. In this population, neither further defibrillation or antiarrhythmic drugs, such as amiodarone and lidocaine, have been shown to improve survival to hospital discharge or neurologically preserved survival [9]. Alternative defibrillation strategies (ADS), i.e. double sequential external defibrillation (DSED) and vector-change defibrillation (VCD), have been suggested for rVF despite the body of evidence is still scanty [10–11]. DSED is the simultaneous use of two sets of manual defibrillators in two different planes (anterior-lateral and anterior-posterior) to double the defibrillation energy [12], while VCD consists in shifting defibrillation pads from the usual anterior-lateral position to the anterior-posterior one [13] (Figure 1). The association between ADS and rVF survival has been studied with discordant findings mainly due to methodological drawbacks of the conducted studies [8,14–22]. Indeed, only two studies reported a significant difference between ADS and standard defibrillation (SD), suggesting a superiority of the former on survival [8,22]. Since survival to hospital discharge might be burdened by several confounding factors related to hospitalization, the present systematic review and meta-analysis aimed to investigate whether ADS improve survival to hospital admission *vs*. SD in adults with non-traumatic rVF.

**Figure 1.**
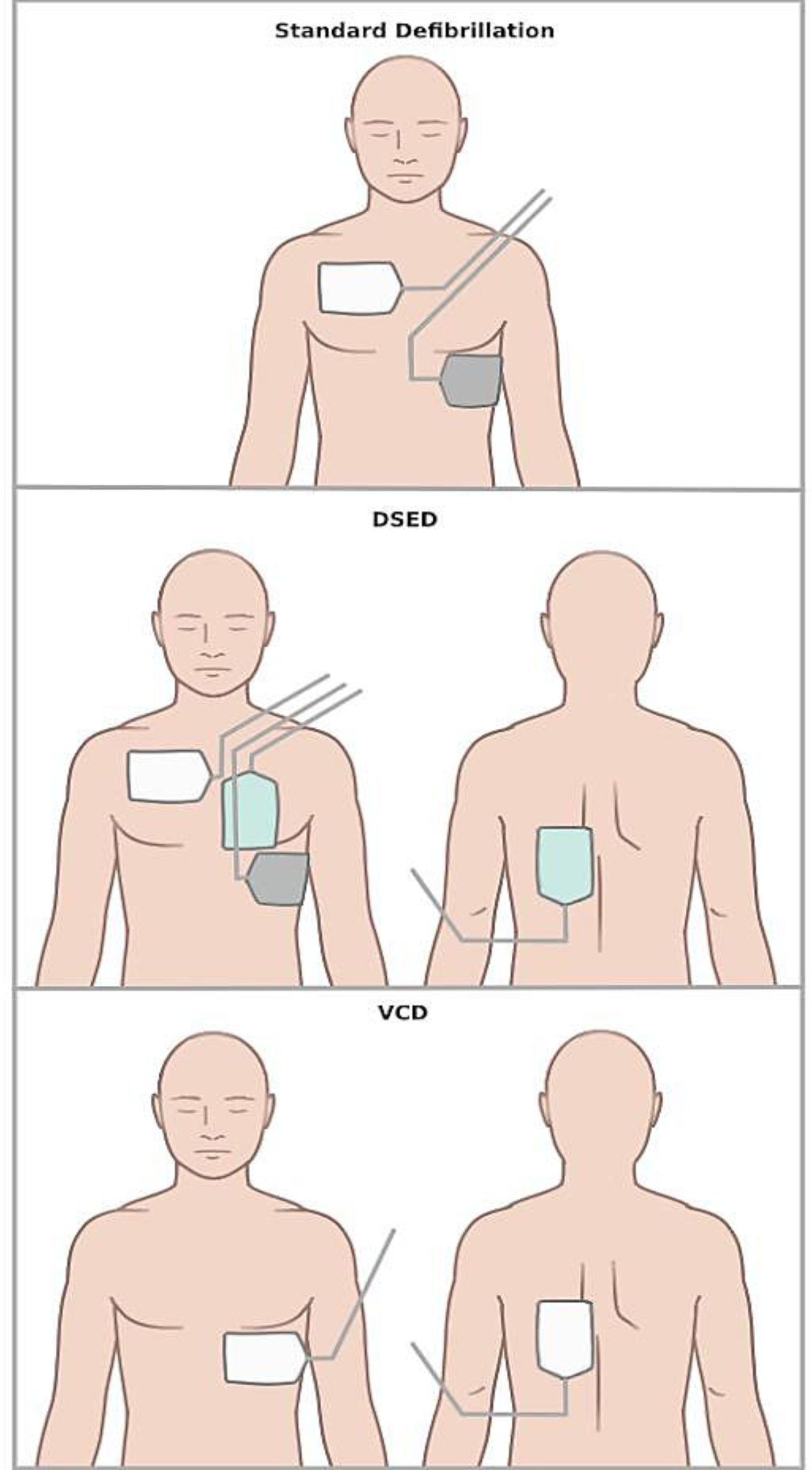
Scheme illustrating pad placement in the different defibrillation strategies. *Notes*. DSED: double sequential external defibrillation; VCD: vector-change defibrillation.

## Methods

This paper has been performed following the Preferred Reporting Items for Systematic Reviews and Meta-Analyses (PRISMA) [23] and Meta-analysis of Observational Studies in Epidemiology (MOOSE) guidelines and checklists [24]. The protocol was registered on PROSPERO (CRD42022379049).

### Data sources and searches

English-language studies on human subjects from MEDLINE, Google Scholar, World Health Organization (WHO), LitCovid NLM, EMBASE, CINAHL Plus and Cochrane Library were retrieved from January 2000 to December 2022. A combination of subject headings and keywords relevant to ADS and rVF were combined using Boolean terms, as appropriate. The complete search strategy included: “refractory ventricular fibrillation” OR “cardiac arrest” OR “ventricular fibrillation” OR “VF” OR “pulseless ventricular tachycardia” OR “pVT” AND “double-sequential external defibrillation” OR “dual-sequential external defibrillation” OR “DSED” OR “vector-change defibrillation” OR “VCD” OR “defibrillation strategies” (see Supplementary Table 1). Furthermore, relevant papers were hand-retrieved to identify further studies that might have been missed by the electronic analysis (BP and MG).

### Study selection

Randomized controlled trials (RCTs), prospective and retrospective studies (including cross-sectional studies and case series with at least five patients) were searched. Studies were eligible if: (1) they compared the use of an ADS *vs*. SD in patients with rVF; (2) patients enrolled were older than 18 years; (3) data on survival to hospital admission were available. Four Authors (RdF, LE, AP and FR), working in pairs, independently screened citation titles and abstracts and retrieved full manuscripts and appendices of potentially relevant articles. Disagreements and inconsistencies were resolved by adjudication with three investigators (BP, MDS, MG).

The meta-analysis was performed comparing studies which expressed the number of patients survived to hospital admission and the sample size of the following subgroups: experimental group (i.e. ADS) *vs.* control group (SD).

### Data extraction and quality assessment

Two reviewers (BP and MG) independently abstracted and recorded data, using standardized data abstraction form (Excel spreadsheet). The investigators were blinded to each other decisions. Disagreements were resolved by consensus and arbitration with a third reviewer (MDS). The following data were extracted: author’s name; year of publication; duration of the analysis; study design; sample size; mean age; female sex; ADS technique; numerosity of experimental and control groups; p-value of ADS *vs.* SD on survival to hospital admission in the univariate analysis; percentage of patients in the experimental group survived to hospital admission; main conclusion. Investigators performed a single contact attempt with study Authors if any data were not available. The quality assessment of the included studies has been performed following the National Institutes of Health (NIH) Quality Assessment Tool for Observational Cohort and Cross-Sectional Studies [25] and the revised Cochrane risk of bias tool for randomized trials (Risk of Bias-2, RoB-2), as appropriate [26]. Two independent investigators scored each article for quality and scoring inconsistencies were resolved by discussion and consensus between the two reviewers (BP and MG).

### Data synthesis and analysis

A random-effects meta-analysis was performed using the *meta* package of R statistical software (version 4.2.2) to estimate the pooled Odds Ratio (OR) with 95% Confidence Interval (95% CI) of the different defibrillation strategies and survival to hospital admission [27]. I² statistic was used to evaluate the heterogeneity across the studies: a considerable heterogeneity was established with an I² statistic >75% and the statistical significance was set at *p* <0.05. All the mentioned analyses were repeated solely on the observational studies, after excluding RCTs.

The publication bias of the involved studies was evaluated both graphically and numerically through Funnel plots and Egger’s regression test, respectively. Two subgroup analyses were performed to compare SD with the single alternative procedures (i.e. DSED or VCD).

## Results

### Literature Search Results

A total of 3360 papers were retrieved through database searching (Figure 2). Hand searching of literature did not identify any further eligible articles. After removing duplicate records, studies conducted on animals and non-English manuscript, 552 papers were identified for screening. Subsequently, 281 citations were excluded, leaving 271 manuscripts for full review. After exclusion of irretrievable papers / studies with unclear or different primary outcome, 8 documents were included in the analysis (Figure 2).

**Figure 2.**
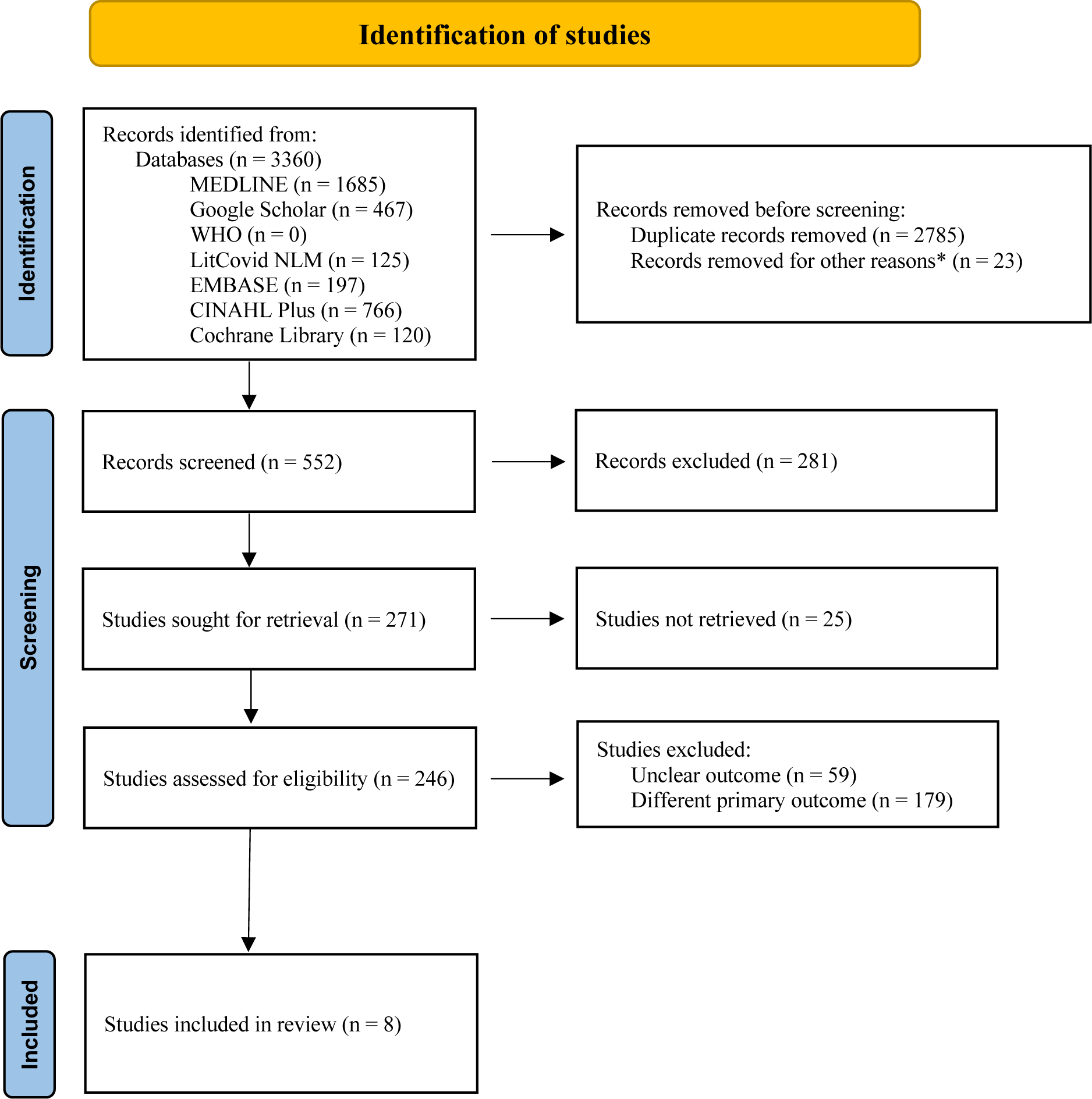
PRISMA 2020 flow diagram for new systematic reviews, which included searches of databases and registers only. *Only human subject-based papers; analysis not in English papers.

### Study characteristics

We retrieved 2 RCTs, 5 retrospective observational and 1 case-control studies. Excluding 152 patients enrolled in both the RCTs by Cheskes *et al*. [8,21], a total of 1405 patients were analysed with a pooled mean age of 61.9 ± 1.1 years. Among participants, 277 (19.7%) were female. Five papers analysed the effect of DSED on survival to hospital admission [16,17,19,20,22], one evaluated VCD *vs*. SD [18] and two RCTs compared ADS to SD in rVF [8,21]. Pooled survival rate in experimental group was 32.3 ± 12.7%. The main features of the selected papers were summarized in Table 1.

**Table 1.** Synopsis of the main features described in papers (n = 8) correlating ADS and survival.

| Author's name | Year | Duration (months) | Study Design | Patients (n) | Mean age (ys) | Female (n) | Type of ASD | Experimental group (n) | Control group (n) | *p* | Survival to hospital admission with ADS (%) | Main conclusion |
| --- | --- | --- | --- | --- | --- | --- | --- | --- | --- | --- | --- | --- |
| Ross *et al* (16) | 2016 | 36 | Retrospective analysis | 279 | 61.0 | 73 | DSED | 50 | 229 | 0.830 | 38.5 | DSED in OHCA did not improve neurologically preserved survival |
| Emmerson *et al* (17) | 2017 | 18 | Retrospective analysis | 220 | 61.9 | 34 | DSED | 45 | 175 | N/A | 30.0 | No clear benefit of DSED vs. SD in rVF |
| Davis *et al* (18) | 2018 | 15 | Retrospective analysis | 25 | 63.0 | 3 | VCD | 16 | 9 | N/A | 37.5 | VCD in rVF may result in VF termination |
| Beck *et al* (19) | 2019 | 48 | Retrospective analysis | 310 | 62 | 75 | DSED | 71 | 239 | N/A | 35.2 | DSED was not associated with OHCA outcomes and may be not beneficial in rVF |
| Mapp *et al* (20) | 2019 | 36 | Case-control study | 128 | 58.4 | 22 | DSED | 25 | 103 | 0.590 | 48.0 | DSED was not associated with improved survival to hospital admission |
| Cheskes *et al* (21) | 2020 | 18 | RCT | 152 | 64.0 | 19 | DSED<br>VCD | DSED: 55<br>VCD: 61 | 36 | N/A | DSED: 32.7<br>VCD: 24.6 | Rates of rVF and ROSC were higher in ADS vs. SD |
| Kim *et al* (22) | 2020 | 24 | Retrospective analysis | 38 | 62.8 | 7 | DSED | 17 | 21 | 0.001 | 82.4 | DSED increased survival to hospital admission vs. SD |
| Cheskes *et al* (8) | 2022 | 50 | RCT | 405 | 63.6 | 63 | DSED<br>VCD | DSED: 125<br>VCD: 144 | 136 | 0.009 | DSED: 30.4<br>VCD: 21.7 | Survival occurred more frequently in ADS |

*Note.* ADS: alternative defibrillation strategies; DSED: double sequential external defibrillation; N/A: not available; OHCA: out-of-hospital cardiac arrest; RCT: randomized controlled trial; ROSC: return of spontaneous circulation; rVF: refractory ventricular fibrillation; SD: standard defibrillation; VCD: vector-change defibrillation.

### Quality assessment and publication bias

Using the NIH quality assessment tool, we evaluated 6 out of 8 studies, as appropriate. Three out of six papers had a poor-quality rating [16–18] and three a fair-quality one [19,20,22]. RoB-2 assessment highlighted an overall low risk of bias of the two RCTs involved [8,21]. No studies were excluded because of a poor-quality rating (<50%). Quality assessments are reported in Supplementary Table 2 and 3. The heterogeneity among all the selected studies was moderate (I² = 62%, p = 0.01) and no publication bias resulted from qualitative (Supplementary Figure 1) and quantitative (Egger regression intercept = 1.029, 95%CI: −2.728, 4.785, p = 0.608) analyses. Among the observational studies, the heterogeneity was low (I² = 27%, p = 0.23) and both the funnel plot and the Egger’s test (Egger regression intercept = 2.223, 95%CI: 0.241, 4.209, p = 0.089) suggested the presence of possible publication bias.

### Defibrillation strategies and survival to hospital admission

The random-effect meta-analysis demonstrated no significant differences in survival to hospital admission among ADS *vs*. SD (n = 1405, pooled OR = 1.12, 95%CI: 0.62, 2.01; Figure 3). This result was confirmed after the exclusion of the patients enrolled in both the RCTs [8,21] (n = 1000, pooled OR=0.86, 95%CI: 0.51, 1.47; Figure 4).

**Figure 3.**
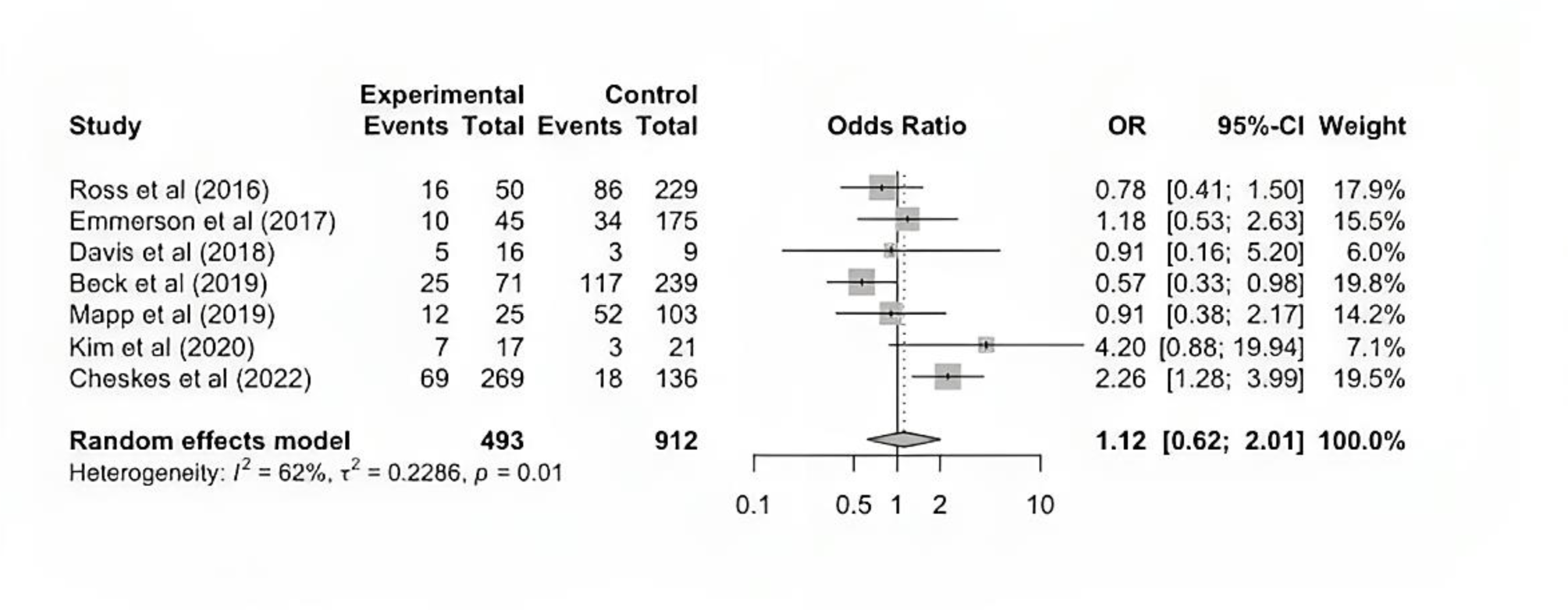
Forest plot on the mean differences of survival to hospital admission between ADS *vs*. SD from the random-effects meta-analysis.

The subgroup analyses confirmed that neither DSED (n=1,236, pooled OR=1.20, 95%CI: 0.56, 2.58; Supplementary Figure 2a) nor VCD (n=305, pooled OR=1.66, 95%CI: 0.10, 27.02; Supplementary Figure 2b) were associated with survival benefit to hospital admission *vs*. SD.

## Discussion

This systematic review and meta-analysis demonstrated that the defibrillation strategy does not influence the rate of survival to hospital admission in adults with rVF.

CA is the most dramatic condition among medical emergencies [6]. Although shockable rhythms have usually better outcomes than non-shockable ones, their progression to rVF is burdened by high mortality rates. The treatment of this condition is still challenging and a specific, well-defined management has not been proposed yet [28]. Even the latest 2021 guidelines by the European Resuscitation Council (ERC) did not conclude for a specific treatment of rVF. Indeed, the Authors suggested to consider the use of VCD, while a weak recommendation against DSED was provided based on low-certainty evidence [29]. Considering these uncertainties and the conflicting data from the RCT recently published [8], the present manuscript aimed at investigating the effects of both DSED and VCD *vs*. SD on survival to hospital admission in patients with rVF.

Most studies included in the systematic review proposed a role of ADS in the management of rVF [8,18,21,22]. Only two studies highlighted a significant difference on survival to hospital admission of ADS *vs*. SD [8,22]. In 2022, Cheskes *et al*. performed a RCT involving a large sample size and showing a low risk of bias at the quality assessment [8]. In contrast, Kim *et al*. carried out a retrospective analysis limited by low numerosity and a questionable quality assessment with an uncommonly high survival rate [22].

Regarding the effect of VCD in rVF, Stupca *et al*. retrospectively assessed 83 patients reporting a negative effect of this technique on return of spontaneous circulation (ROSC) and survival to hospital admission / discharge [30]. However, VCD-treated patients also received active pharmacological therapies (i.e., esmolol and dose-capped epinephrine), thus leading to the exclusion of this study from our analysis.

The main message emerged from our analysis was the lack of difference on survival to hospital admission between ADS and SD. This finding is in line with the conclusions of previous systematic review [31] and meta-analyses [32,33]. Nonetheless, our analysis comprehended the most recent RCT [8] and addressed a different primary outcome from the aforementioned studies. Indeed, our main goal was the evaluation of the survival to hospital admission instead of survival to hospital discharge, which may be biased by confounding factors related to hospitalization. The subgroup analyses confirmed that neither DSED nor VCD *vs*. SD improved survival to hospital admission in patients with rVF, corroborating the results obtained in the meta-analysis performed on ADS.

Despite the 2016 Sex And Gender Equity In Research guidelines encouraged to perform a sex- and gender-based analysis (SGBA), none of the included studies reported sex-disaggregated findings [34]. Moreover, although up to 42% of patients experiencing CA were females [4,5], the percentage of women enrolled in the included studies was only 19.0%. The female underrepresentation in clinical studies has been commonly reported, especially in cardiovascular clinical trials [35,36], limiting the generalizability of trial findings. Also in the present meta-analysis, we confirmed the lack of data on treatment outcomes in female adults with rVF. Recently, sex-based differences in CA management and survival have been reported. Female adults experiencing OHCA had a small absolute difference for the outcome survival to discharge as compared with the male counterpart, while no difference in survival at 30 days was detected [37]. A systematic review also reported how the unadjusted difference in the outcomes of male and female with CA may be explained by the worse phenotype of women such as the older age, greater burden of comorbidities and the higher proportion of less witnessed arrests with no shockable rhythms [38]. Therefore, more studies are needed to improve our understanding of CA in women.

The main strengths of the present analysis are that: i) it was the first systematic review and meta-analysis highlighting the effect of the different type of ADS on survival to hospital admission; it was focused on a clinically meaningful outcome, influenced mostly by the applied treatment; the heterogeneity of the included studies was moderate and no publication bias was detected. Our findings should be carefully interpretated in the light of several limitations. First, inclusion criteria were met only by few studies and, among them, only two were RCTs. Second, the overall sample size exhibited a scarce numerosity with an unbalanced representation of female adults and no SGBA performed. Third, we considered only survival to hospital admission which, although clinically meaningful, did not describe the functional status of the discharged patients.

In conclusion, this systematic review and meta-analysis, which included the latest and largest RCT comparing the use of ADS *vs.* SD, did not show any difference on survival to hospital admission between the different defibrillation techniques in rVF. We acknowledge the statistical strength of a well-conducted RCT demonstrating the survival benefit of ADS *vs*. SD. Nevertheless, considering the contrasting results emerged from this meta-analysis, our study advocates for further well-designed clinical trials, including SGBA, aimed at establishing the actual role of ADS.

## Data Availability

All data and statistical analyses codes are available upon request to the corresponding author.

## Acknowledgments

The authors wish to thank Dr. Donato Bragatto, Dr. Claudia Righini, Mrs. Manuela Zappaterra (Biblioteca Interaziendale di Scienza della Salute, Hospital of Ferrara), Mrs. Egizia Zironi and Mrs. Silvia Bellotti (Unità Servizi Interbibliotecari, University of Ferrara) for their valuable collaboration. Furthermore, the authors thank Alice Eleonora Cesaro and Greta Francesca Cesaro for image editing.

## Abbreviations

ADS: alternative defibrillation strategies
CA: cardiac arrest
DSED: double sequential external defibrillation
EMS: emergency medical service
ERC: European Resuscitation Council
IHCA: in-hospital cardiac arrest
MOOSE: meta-analysis of observational studies in epidemiology
OHCA: out-of-hospital cardiac arrest
NIH: National Institutes of Health
PEA: pulseless electric activity
PRISMA: preferred reporting items for systematic reviews and meta-analyses
pVT: pulseless ventricular tachycardia
RCT: randomized controlled trial
RoB-2: Risk of Bias-2
ROSC: return of spontaneous circulation
rVF: refractory ventricular fibrillation
SD: standard defibrillation
SGBA: sex- and gender-based analysis
VCD: vector-change defibrillation
VF: ventricular fibrillation.

## Statements and Declaration

### Competing interests

Authors have no conflicts of interest to disclose.

### Funding

RDG was supported by FAR (Fondo Ateneo Ricerca) and FIR (Fondo Incentivazione Ricerca) funds from the University of Ferrara, Italy.

### Reproducible Research Statement

The study protocol was registered on PROSPERO (CRD42022379049).

### Statistical code and data set

All data and statistical analyses codes are available upon request to the corresponding author.

### Ethical Approval

This research does not directly involve patients; hence, an ethical approval was deemed unnecessary.

### Authors’ contributions

BP and MG designed the project and wrote the paper. BP, MG, RdF, LE, AP and FR built the database. FRe and CT analysed the database. BP, MG, VR, GM, GB, VS, SP, MV, MDS and RDG critically reviewed the paper. All authors have read and agreed to the published version of the manuscript.

### Availability of data and materials

The dataset is available for reviewers on reasonable request.

---

**Supplementary Figure 1.**
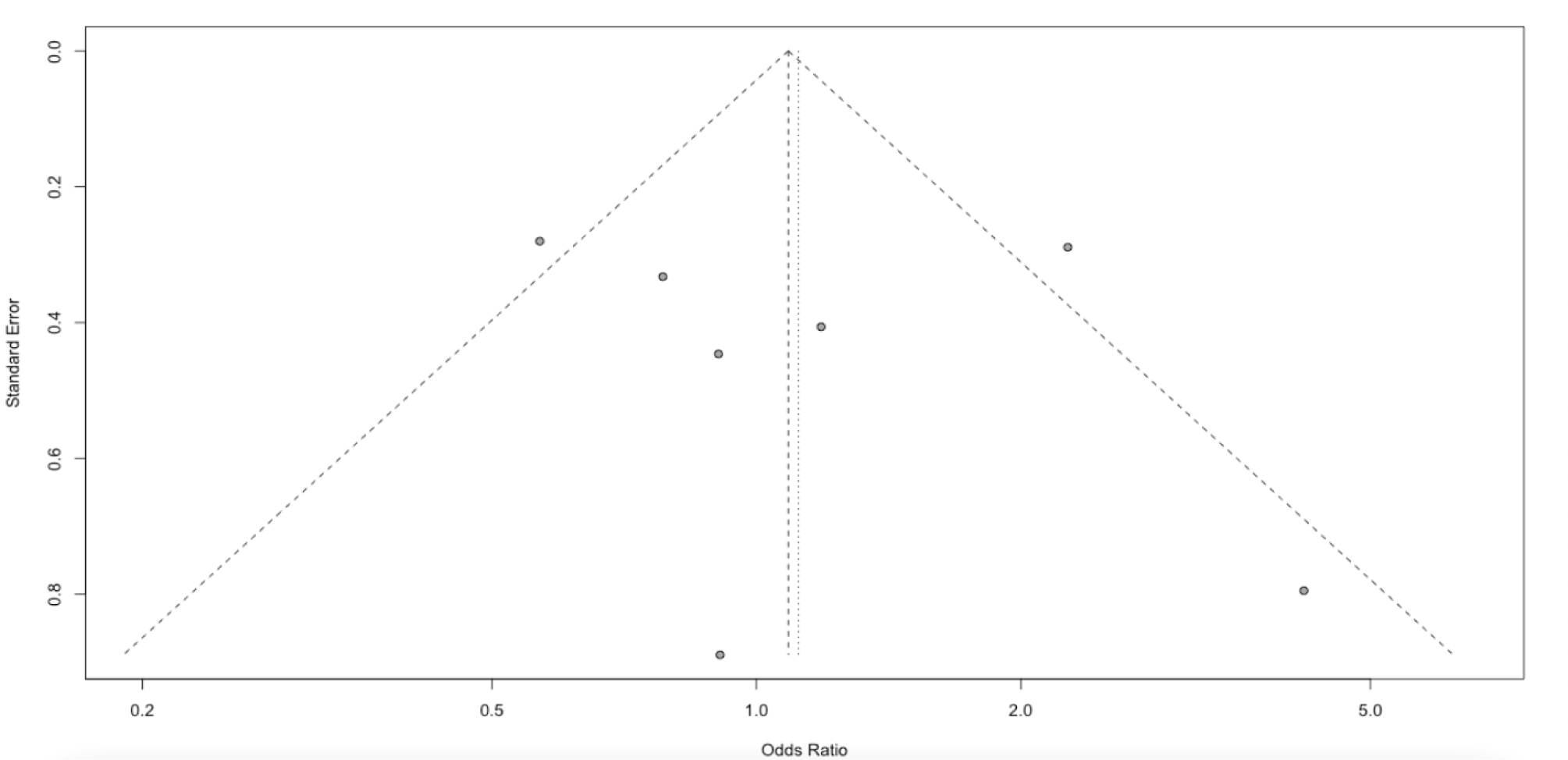
Funnel plot of the included studies.

**Supplementary Figure 2.**
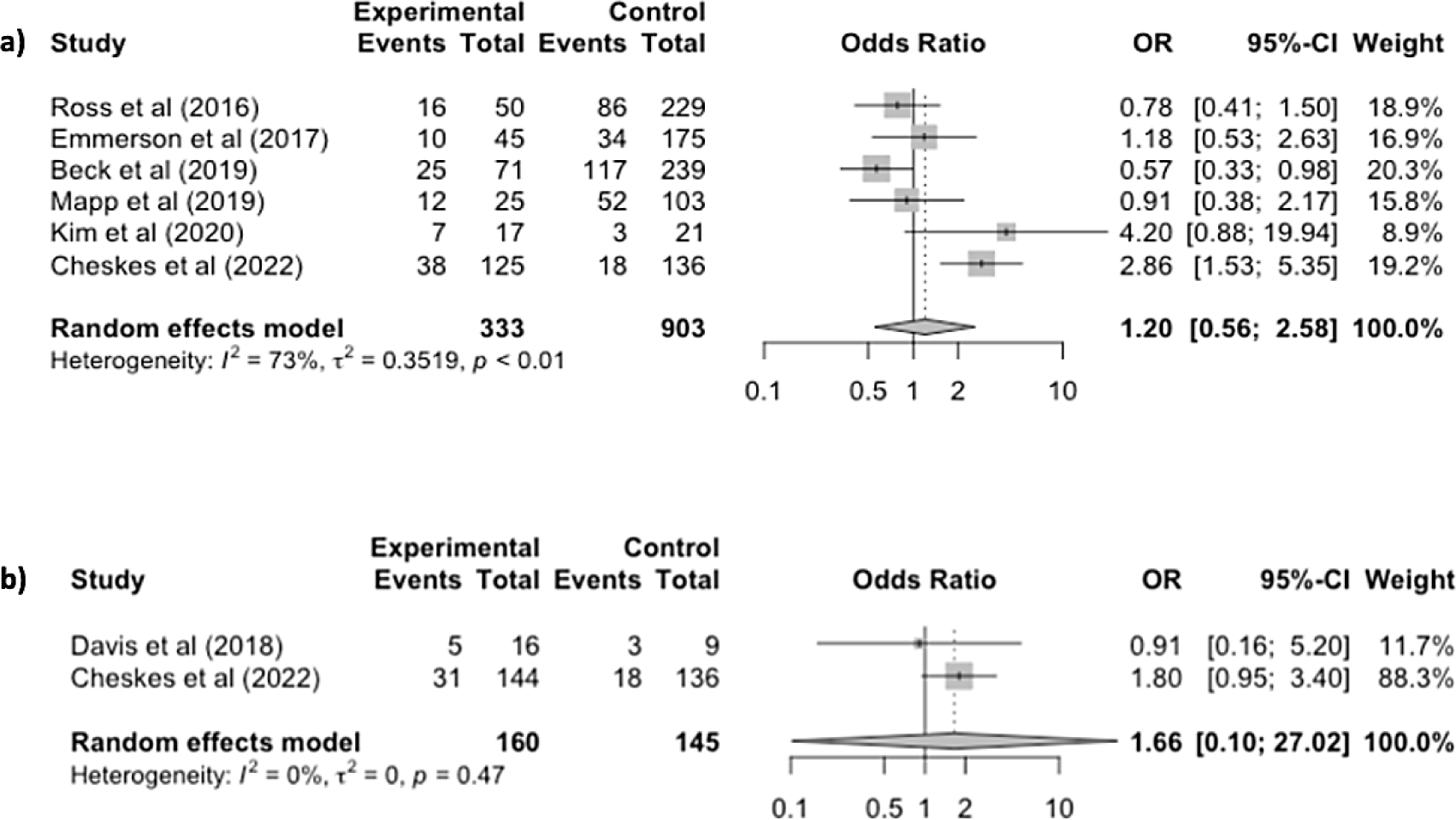
Forest plot on the mean differences of survival to hospital admission between DSED (2a) / VCD (2b) *vs*. SD from subgroup analysis.

**Supplementary Table 1.**
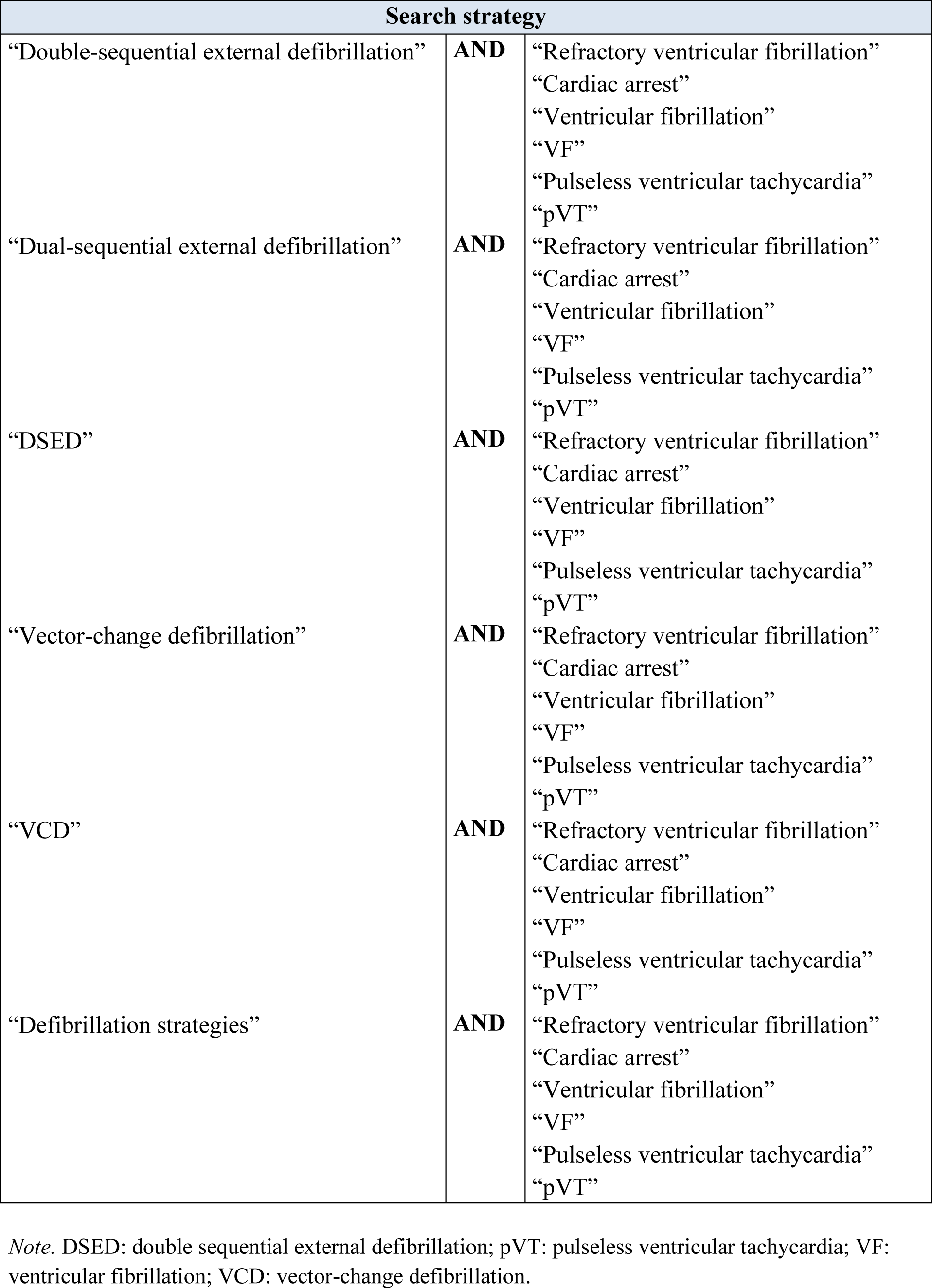
Complete literature search strategy.

**Supplementary Table 2.**
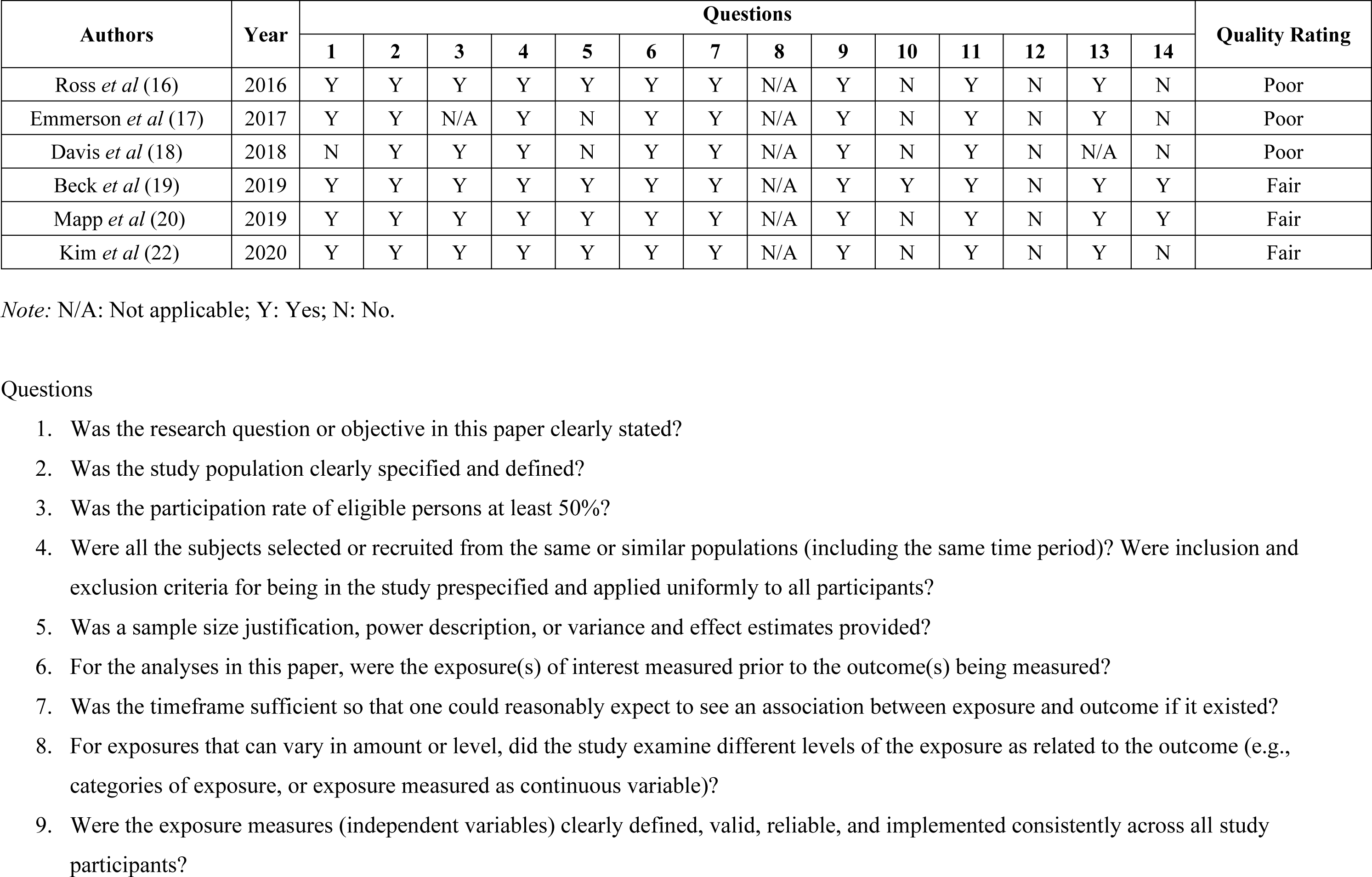
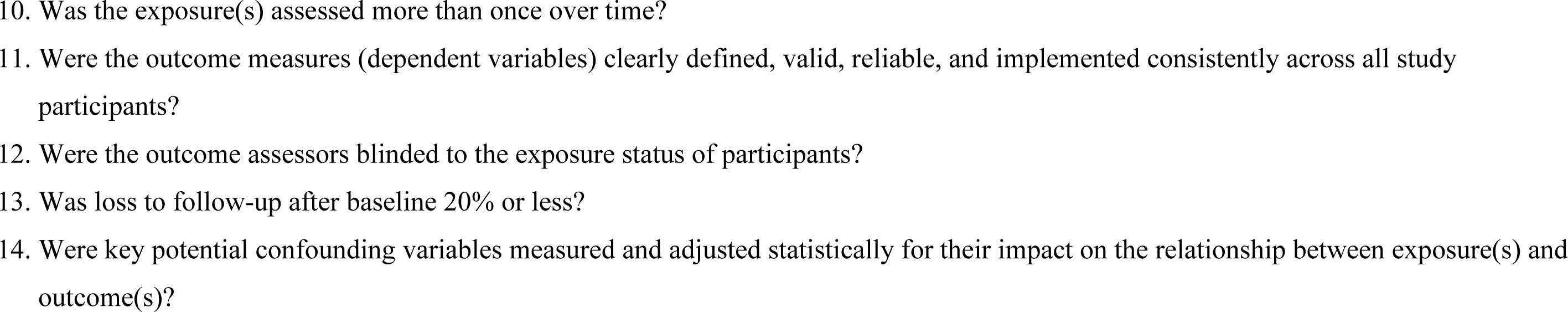
NIH Quality Assessment Tool for Observational Cohort and Cross-Sectional Studies.

**Supplementary Table 3.**
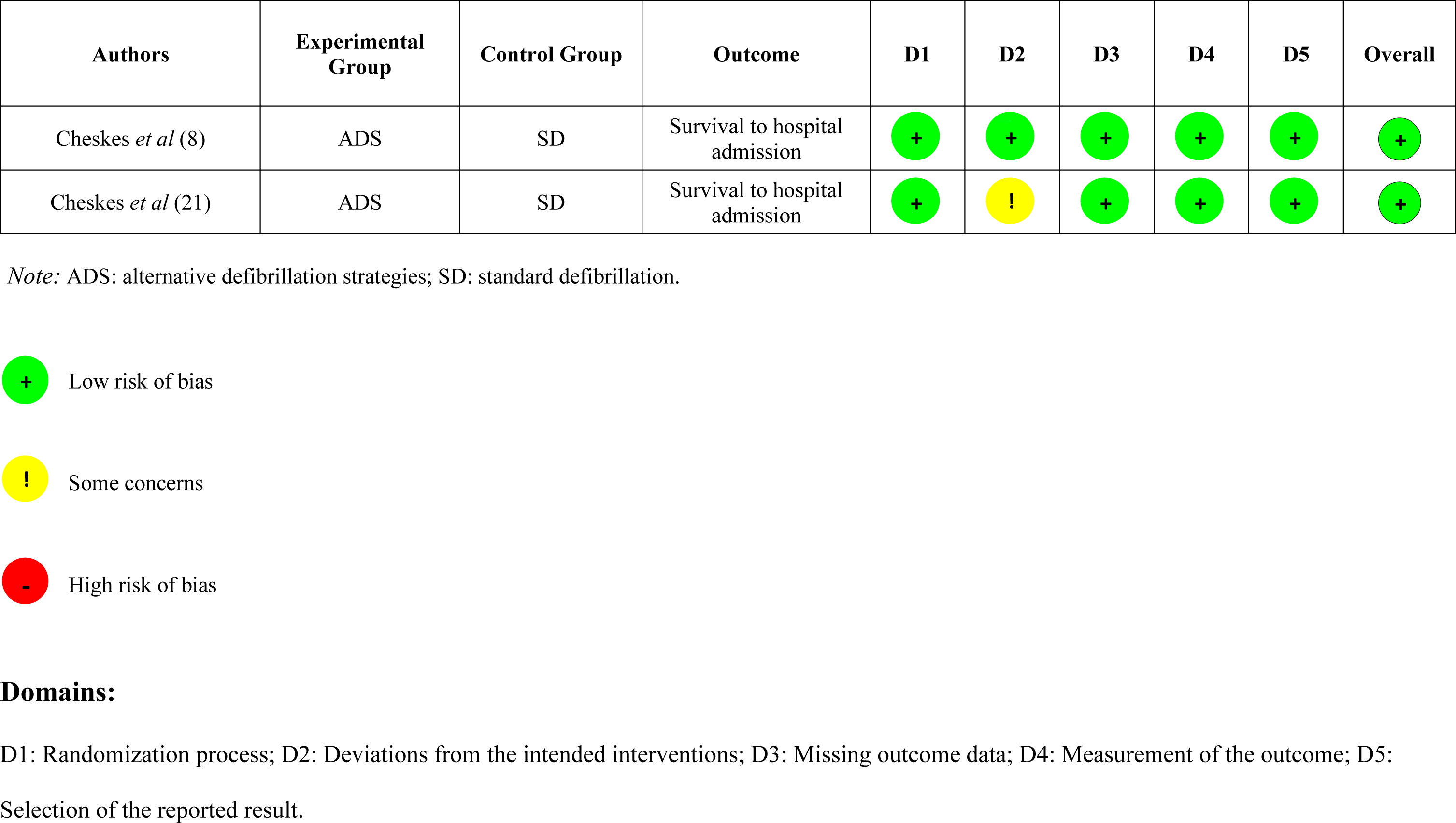
Risk of Bias-2 (RoB-2) assessment for randomized trials.

## Notes

### Competing Interest Statement

The authors have declared no competing interest.

### Author Declarations

The study protocol was registered on PROSPERO (CRD42022379049). This research does not directly involve patients; hence, an ethical approval was deemed unnecessary.

